# Predicting Future Brain Atrophy Based on Longitudinal MRI

**DOI:** 10.1101/2025.04.09.25325520

**Authors:** Maryam Hadji, Elaheh Moradi, Jussi Tohka

## Abstract

Neuron loss is a key feature of neurodegenerative diseases often leading to brain atrophy detectable through magnetic resonance imaging (MRI). Various brain atrophy measures are essential in research of Alzheimer’ disease (AD) and related dementias. This study aims to forecast future annual percentage changes in hippocampal, ventricular, and total gray matter (TGM) volumes in individuals with varying cognitive statuses, from healthy to dementia. We developed a machine learning model using elastic net linear regression and tested two approaches: (1) a baseline model using predictors from a single-time-point and (2) a longitudinal model using predictors derived from longitudinal MRI. Both approaches were evaluated with MRI-only models and models that combined MRI with additional risk factors (age, sex, APOE4, and baseline diagnosis). Cross-validated Pearson correlation scores between predicted and actual annual percentage changes were 0.62 for the hippocampus, 0.51 for the ventricles, and 0.41 for TGM, using the longitudinal MRI + risk factor model. Longitudinal models consistently outperformed baseline models, and models including risk factors outperformed the MRI only model. Validation using an external dataset confirmed these findings, highlighting the value of predictors derived based on longitudinal data. We further studied the value of the predicted atrophy/enlargement rates for clinical status progression prediction across three different datasets. Predicted atrophy was a consistently better indicator of progression to mild cognitive impairment and dementia than present-day regional volumes, with the longitudinal atrophy prediction model typically outperforming the baseline model in terms of clinical status prediction. Future atrophy prediction has significant potential for assessing the risk of cognitive decline, even in cognitively unimpaired individuals, and can aid in selecting participants for clinical trials of disease-modifying drugs for AD.

## 1 Introduction

Dementia impacts approximately 57 million people globally [1], with Alzheimer’s disease (AD) being its most common cause [2]. Machine learning approaches for predicting future diagnoses have been extensively studied in the context of dementia [3], with numerous studies focusing on the progression from mild cognitive impairment (MCI) to dementia [4]. However, as the clinical focus shifts from prognosis to selecting the most effective treatments, there is a growing need to predict more nuanced outcomes [5, 6]. This shift in focus could also enhance the selection of study participants for clinical trials, ensuring that the most appropriate candidates are chosen based on their predicted responses to treatment.

Predicting the change in cognitive tests scores, such as mini mental state examination (MMSE) and Alzheimer’s Disease Assessment Scale–Cognitive Subscale (ADAS-Cog), is perhaps the most natural way towards these more nuanced outcomes. However, predicting changes in cognitive scores has proven to be challenging, partly due to the validity challenges in the longitudinal measurement of cognitive changes in less impaired populations with the well-established tests [7, 8] and partly due to challenges in assessing cognition in general [9]. These challenges include learning effects, which can skew results due to repeated testing, and high variability, where outcomes depend significantly on the individual’s condition on the test day. For example, in the recent TADPOLE challenge, none of the competing teams was able to perform better than a simple baseline method for the ADAS13 score prediction while the predictions for the future diagnosis and future ventricular volume were significantly better than with the baseline methods [10]. Moreover, while the future cognitive scores may be predictable, this predictive power is many times driven by the current baseline scores, yielding change estimates of limited usability [11]. As a result, there is a pressing need for serial biomarkers that reflect the underlying biology of AD and its connection to dementia.

This work, instead, explores the prediction of future atrophy based on serial magnetic resonance imaging (MRI). The MRI-based brain atrophy has long been identified as an important measure to track dementia progression [12]. It possesses several important characteristics for a prediction target: 1) atrophy measures are quantitative, 2) MRI is widely available and non-invasive, 3) various atrophy measures have been adopted as secondary endpoints in randomized controlled trials of dementia/AD interventions [13, 14] and 4) hippocampal atrophy has been qualified for enrichment in pre-dementia trials by the European Medicines Agency [15]. By focusing on atrophy prediction, this study aims to provide a more reliable and clinically useful tool for both treatment selection and trial participant selection.

We focus on three distinct atrophy measures: hippocampal atrophy, enlarged ventricular volume, and whole-brain atrophy which all have been established as key structural MRI markers for AD and dementia progression, with slightly different time windows of the best predictive utility [16–19]. In clinical trials on dementia and AD, the atrophy of these three key brain regions are often considered as secondary endpoints (e.g. [20]). These regions provide valuable insights into short-term dementia risks and changes in their volumes has been linked with the cognition [21–23].

Surprisingly, while the association of brain atrophy measures to dementia and AD progression is well documented, the prediction of future changes in these measures has not been studied widely. Prediction of the future ventricle volume was one of the tasks in a recent TADPOLE challenge [10], where the best performing model was based on the concept of rate of AD progression [24]. Liedes et al [25] focused on prediction of hippocampal atrophy with promising results. However, in [25] they studied only prediction of hippocampal atrophy and used data from single time-points only for predictions. In this work, we analyze three regions (hippocampus, total gray matter, and ventricles) together within the same dataset and workflow and develop a modeling framework able to use predictors derived from longitudinal data. Finally, we study the value of forecasted atrophy rates to predict the progression of the level of the memory concern from healthy to MCI/dementia and from MCI to dementia. We demonstrate that the models that use predictors derived from longitudinal data consistently outperform corresponding models based on single time-point predictors. Also, we show that predicted atrophy is a better indicator of progression to mild cognitive impairment and dementia than the volumes of the regions at current day.

## 2 Materials and Methods

### 2.1 Data for atrophy/enlargement prediction

Data used in this work were obtained from the ADNI (http://adni.loni.usc.edu) and the AIBL study group (https://aibl.org.au). The ADNI was launched in 2003 as a public-private partnership, led by Principal Investigator Michael W. Weiner, MD. The primary goal has been to test whether serial MRI, PET, other biological markers, and clinical and neuropsychological assessment can be combined to measure the progression of MCI and early AD. For up-to-date information, see (www.adni-info.org). AIBL study methodology has been reported previously [26].

We included all ADNI participants who had a T1-weighted MRI acquired at the time of entry, 24-month visit, and 48-month visit. Moreover, we required participants to have information on demographics (age, sex), APOE4 status, and diagnosis status obtained from from the ADNIMERGE.csv table. 2,365 participants had MRI data at the time of entry. After 24 months of follow-up, MRI information was available for 1,373 participants, and for 586 participants after 48 months. 565 individuals had MRI data at entry, 24 months, and 48 months. After excluding participants without APOE4 information, we finalized our dataset with 550 individuals who had all the considered information. We refer to this dataset as ADNI-main dataset.

We used the Australian Imaging Biomarkers and Lifestyle Flagship Study of Aging (AIBL, ADNI-subcohort) as a external test data in our study. At the study entry, MRI scans were available for 605 participants. After 18 months of follow-up, 232 individuals had MRI scans, and 149 individuals had scans at the 36-month follow-up. Among them, 95 participants had MRI scans at all three time points. All these 95 individuals have complete demographic data, including age, sex, diagnosis, and APOE4 status. The characteristics of study cohorts are presented in Table 1 and participant’s RIDs are available as supplementary material.

**Table 1.** Characteristics of the ADNI and AIBL cohorts, including sample size, age, sex distribution, APOE4 status, and clinical diagnosis. Age is presented as mean (standard deviation). Diagnosis and age are from the time of entry. CN refers to cognitively normal. APOE4 status is displayed as the count of individuals with 0, 1, or 2 alleles.

| Baseline characteristics | ADNI | AIBL |
| --- | --- | --- |
| Sample size, N | 550 | 95 |
| Age, years | 73.04 (6.07) | 73.31 (6.93) |
| Sex, M/F | 297 / 253 | 48 / 47 |
| APOE4, 0/1/2 | 321 / 186 / 43 | 60 / 29 / 6 |
| Diagnosis, CN/MCI/AD | 246 / 236 / 68 | 76 / 12 / 7 |

### 2.2 MRI preprocessing

The MRI data were preprocessed following the framework described in [27]. We used the volumetric and thickness measures computed with CAT12 toolbox (https://neuro-jena.github.io/cat/, version 8.1) using the default settings [28]: For 136 volume measures, the T1-weighted MRI was segmented into gray matter (GM) and white matter (WM) and non-linearly normalized to a stereotactic space using the shooting approach [29]. Based on spatially normalized GM and WM segments, the neuromorphometrics atlas was used to extract the regional volumes^1^. We denote these volume measures by *V*_*ij*_(*t*) for participant *i*, region *j* = 1, …, 136, and time point *t*. The cortical thickness measures [30] were computed and registered to the FSaverage surface template [31], where DK40 atlas [32] was used to define 69 regionally averaged cortical thickness measurements. These cortical thickness measures are denoted by *V*_*ij*_(*t*) for participant *i*, region *j* = 137 …, 206, and time point *t*. The MRI data from the participant *i* at time point *t* is collected into a vector *V*_*i*_(*t*).

The annual percentage change between the time points *t*_1_ and *t*_2_ (expressed in years) for a region *i* is defined as

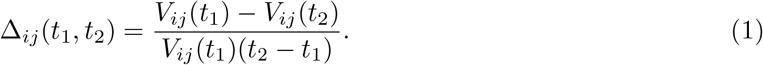

If Δ_*ij*_(*t*_1_, *t*_2_) is positive, it refers to atrophy, and if Δ_*ij*_(*t*_1_, *t*_2_) is negative, it refers to enlargement.

### 2.3 Prediction framework

Our purpose is to predict future Δ_*ik*_(*t*_2_, *t*_3_) based on the data available during time points *t*_1_ and *t*_2_ where *t*_3_ *> t*_2_ *> t*_1_. For simplifying the terminology, we name the time-point *t*_1_ as past, *t*_2_ as current day, and *t*_3_ as future. We formulate this prediction as supervised learning problem. Denote future change Δ_*ik*_(*t*_2_, *t*_3_) by *y*_*ik*_ and the number of participants by *N*. Provided the data **x**_*i*_, *i* = 1, …, *N* (see Table 2) from the past and current day, we apply the Elastic Net linear regression, a regularization technique that combines both Lasso (L1) and Ridge penalties (L2) [33]. Elastic Net is advantageous in high-dimensional data environments, especially when multicollinearity (correlation among predictors) is present. This method allows us to improve model generalization by reducing over-fitting while selecting a sparse subset of informative features. Dropping the index *k*, elastic net solves the coefficients 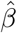 of the linear model as

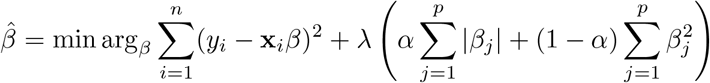

where *β* = [*β*_1_, …, *β*_*p*_]^*T*^, *λ* controls the overall regularization strength, and *α* adjusts the balance between L1 and L2 penalties. When *α* = 1, Elastic Net behaves like Lasso regression, driving some coefficients (*β*_*j*_) to exactly zero, effectively selecting relevant features. Conversely, when *α* = 0, it acts as Ridge regression, reducing the effect of multicollinearity without eliminating predictors. By setting 0 *< α <* 1, Elastic Net captures the advantages of both Lasso and Ridge regularization, yielding a stable and interpretable model even in the presence of correlated predictors.

**Table 2.**
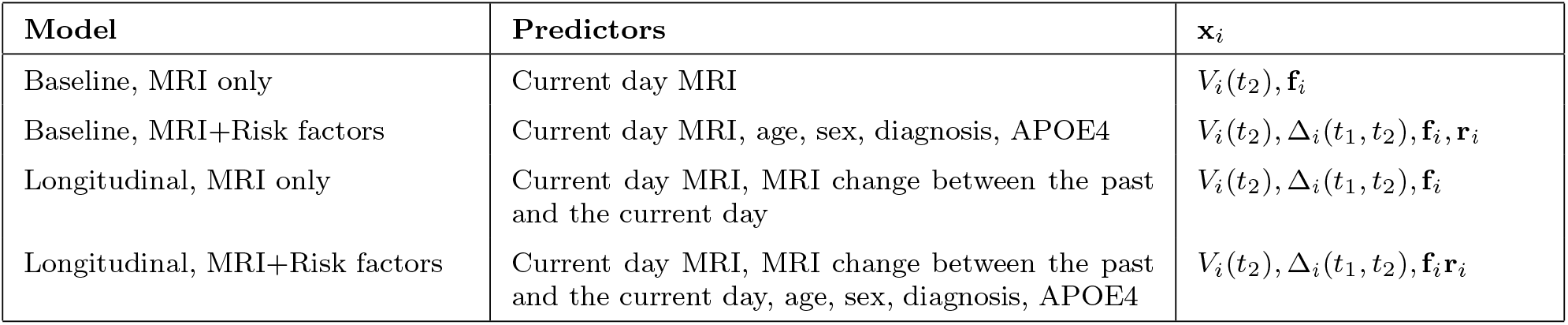
Definitions of the predictive models used in the study. The models vary by the inclusion of MRI data, MRI changes over time, and additional risk factors such as age, sex, diagnosis, and APOE4 status.

| Model | Predictors | $\mathbf{x}_i$ |
| --- | --- | --- |
| Baseline, MRI only | Current day MRI | $V_i(t_2), \mathbf{f}_i$ |
| Baseline, MRI+Risk factors | Current day MRI, age, sex, diagnosis, APOE4 | $V_i(t_2), \Delta_i(t_1, t_2), \mathbf{f}_i, \mathbf{r}_i$ |
| Longitudinal, MRI only | Current day MRI, MRI change between the past and the current day | $V_i(t_2), \Delta_i(t_1, t_2), \mathbf{f}_i$ |
| Longitudinal, MRI+Risk factors | Current day MRI, MRI change between the past and the current day, age, sex, diagnosis, APOE4 | $V_i(t_2), \Delta_i(t_1, t_2), \mathbf{f}_i, \mathbf{r}_i$ |

We used (nested) cross-validation to optimize the parameter *λ*, ensuring that the selected model provided the best trade-off between model complexity and predictive accuracy. The parameter *α* was fixed at 0.5 following our previous works [34]. This approach allowed us to obtain a model that is both interpretable and well-suited for generalization to new data.

We performed the prediction for three different regions of interest: 1) hippocampus, defined as the union of left and right hippocampus; 2) ventricles, defined as the union of left and right lateral ventricles and inferior lateral ventricles, and 3) total gray matter (TGM) volume. Hippocampus and ventricles were defined according to the neuromorphometrics atlas (https://neuromorphometrics.com/Seg/html/segmentation/) and the TGM volume was defined as in the CAT12 toolbox [28].

We designed four models based on two approaches with different feature combinations, presented in Table 2. In the baseline approach, in which we used current day data (ADNI 24 month visits) to predict atrophy in the future (ADNI 48 month visits), we defined two models: the MRI only model was trained using only MRI information and the MRI and risk factors model used in addition demographic/risk factor information containing age, sex, the number of APOE4 alleles, and the current day diagnosis (normal cognition, MCI, dementia). We represented this demographic/risk-factor information by **r**_*i*_ for participant *i*. The longitudinal approach used past and current day data (ADNI 0 and 24 month visits) to predict atrophy in the future (ADNI 48 month visits), and we again defined MRI only and MRI+ risk factors models. Since the MRIs were acquired both with 1.5T and 3T scanners, we provided this information to the learning algorithm as additional predictors (see [35]), encoding the field-strength combination for participant *i* as a binary vector **f**_*i*_.

### 2.4 Implementation and performance evaluation

Following the framework described in [36], we implemented a 10-fold cross-validation (CV) approach to prevent overfitting and evaluate model performance, see Figure 1. The data was divided into training and test sets using a nested and stratified 10-fold CV framework [37]. This involved two levels of CV: an outer loop for model evaluation and an inner loop for selecting hyperparameters (*λ*). Participants were randomly divided into 10 subsets. In each fold of the outer loop, one subset was used for testing, and the remaining nine were designated for training. During the inner loop, optimal *λ* were selected by minimizing the mean square error (MSE). This nested approach ensures robust model evaluation and minimizes overfitting by effectively utilizing the entire dataset for training and testing.

**Fig. 1.**
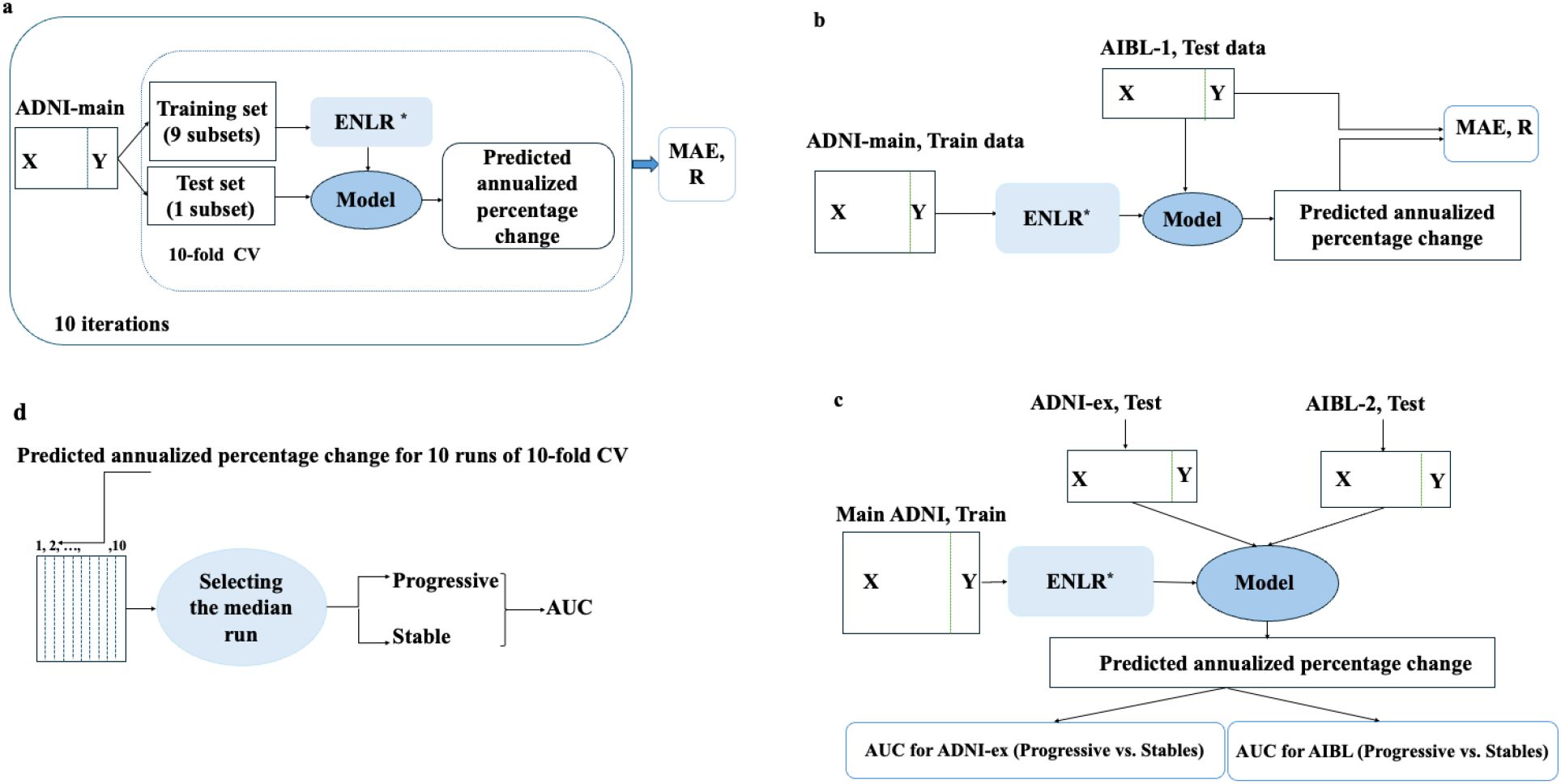
Schematic representation of the prediction framework. **(a)** Atrophy prediction framework: The model is trained and validated using a 10-fold cross-validation approach on the ADNI-main dataset. Elastic Net Linear Regression (ELNR) is used to predict the annualized percentage change. The performance is evaluated using Mean Absolute Error (MAE) and Pearson correlation (R). **(b)** External validation: The trained model is applied to the AIBL-1 test dataset (Table 1) predict the annualized percentage change, and the performance is assessed using MAE and Pearson correlation (R). **(c)** Progression prediction: The model is trained on the ADNI-main dataset and applied to external test datasets (ADNI-ex and AIBL-2, Table 3) to predict clinical progression. The Area Under the Curve (AUC) is used to evaluate classification performance for progressive vs. stable cases. **(d)** Selection of the representative run: The median run is selected from 10 iterations of 10-fold cross-validation, and participants are classified into progressive or stable groups based on predicted values. AUC is then calculated for the classification task. We use AIBL-1 and AIBL-2 to refer to two different versions of the AIBL test data, described in Tables 1 and 3, respectively.

We assessed the models’ performance using the cross-validated Pearson correlation coefficient (PCC) and the mean absolute error (MAE) between the predicted and actual rates of cognitive decline expressed in percentage points. To ensure robustness and minimize the influence of random variations, the results were averaged over 10 runs of nested 10-fold CV. To compare the correlation coefficients between the two approaches, we tested the results of a computation run with median PCC (6th largest PCC among 10 runs) using Hittner-modification of the Dunn-Clark test [38, 39] implemented in the CoCor package [40]. We computed the confidence intervals using a bootstrap procedure specifically designed for repeated CV [41]. We considered all the comparisons planned and therefore report the results of all comparisons without adjustments for multiple testing.

**Table 3.** Demographic and clinical characteristics of cohort used for validation of clinical progression prediction. The cohorts include ADNI-Main, AIBL, and ADNI-External datasets, with subgroups categorized as CN Progressive, CN Stable, MCI Progressive, and MCI Stable. Age is presented as mean (standard deviation). Sex is shown as M/F, and APOE4 status is displayed as the count of individuals with 0, 1, or 2 alleles.

| datasets | Group | N | Age, years | Sex, M/F | APOE4 (0/1/2) |
| --- | --- | --- | --- | --- | --- |
| ADNI-Main | CN Progressive | 60 | 73.74 (6.63) | 32 / 28 | 51 / 8 / 1 |
|  | CN Stable | 133 | 74.71 (5.34) | 64 / 69 | 85 / 46 / 2 |
|  | MCI Progressive | 79 | 71.90 (7.24) | 44 / 35 | 53 / 24 / 2 |
|  | MCI Stable | 119 | 72.64 (7.15) | 71 / 48 | 56 / 46 / 17 |
| AIBL | CN Progressive | 13 | 74.38 (6.56) | 10 / 3 | 4 / 7 / 2 |
|  | CN Stable | 45 | 73.51 (7.07) | 16 / 29 | 36 / 8 / 1 |
| ADNI-External | CN Progressive | 25 | 75.42 (5.27) | 18 / 7 | 13 / 12 / 0 |
|  | CN Stable | 56 | 69.69 (5.74) | 21 / 35 | 42 / 13 / 1 |
|  | MCI Progressive | 46 | 73.81 (6.78) | 29 / 17 | 23 / 17 / 6 |
|  | MCI Stable | 43 | 71.08 (7.56) | 24 / 19 | 30 / 13 / 0 |

Data in the ADNI originated from different phases of ADNI (ADNI1, ADNI2, ADNI3, ADNIGO) with different MRI protocols and scanner field strengths (1.5T or 3.0T). Our baseline approach to correct for differences in MRI markers due to scanner field strength was to include this information in the prediction model as explained in Section 2.3. Additionally, we tested ComBat to harmonize MRI data. ComBat, an empirical Bayes method for batch correction in microarray expression data [42], has been successfully utilized for harmonization in MRI [43]. However, the addition of a harmonization step using the ComBat approach did not improve the prediction performance compared to our baseline approach (Supplementary Table 1).

For ADNI and hippocampal atrophy prediction, we excluded data from two individuals with biologically implausible hippocampal change rate, indicating annual volume increase above 2%. We performed visual quality control on the data from these individuals that indicated poor quality registration or segmentation.

We further utilized the AIBL-cohort as an independent test dataset. The ADNI data served as the training set, while AIBL data was used for testing. The visit schedule of AIBL is different from the ADNI visit schedule: the current day are 18 (AIBL) and 24 (ADNI) month visit, and future are 36 (AIBL) and 48 (ADNI) month visit. However, as the prediction task was formulated as the prediction of annualized change, this causes no differences to the prediction framework presented above. All the analyses were done using R (version 4.1.1), with the following packages: glmnet [44], caret [45], cocor [46], pROC [47], ggplot2 [48], and complexheatmap [49]. The codes to reproduce the work are available at https://github.com/MaryamHadji/Future-prediction

### 2.5 MCI/dementia progression prediction

We evaluated the effectiveness of our estimated atrophy/enlargement rates in predicting dementia progression (Figure 1 c&d). This analysis included participants from the CN and MCI groups. We predicted their future atrophy/enlargement rates and assessed how well these estimates predicted later changes in cognitive status, specifically CN-to-MCI/dementia or MCI-to-dementia progressions. Model’s performance was measured using the area under the receiver operating characteristic curve (AUC), and we considered each region of interest (hippocampus, ventricles, and TGM) separately. We divided participants within the CN and MCI groups into two categories based on their diagnosis at the current visit and follow-up data: 1) Stable group: CN/MCI individuals who have remained with the same diagnosis status for five years or more following the current day visit; 2) Progressive group: a) Individuals with CN diagnosis at the current day visit who later develop MCI or dementia (the last two diagnoses must be MCI or dementia), and b) MCI individuals who later develop to dementia (the last two diagnoses must be dementia). We then compared the ability of predicted future atrophy/enlargement rates to MCI/dementia progression prediction against a baseline using presentday brain region volumes. We hypothesized that predicted atrophy/enlargement rates better reflect future MCI/dementia progression than present-day brain region volume.

We used three different datasets for MCI/Dementia progression prediction: First, we used the main ADNI dataset, which we had previously used for atrophy prediction, comprising data from 550 individuals. Among the 246 CN individuals, 60 individuals progressed MCI/dementia during the followup period, and 133 remained stable with a follow-up duration of at least five years. Similarly, in the MCI group, 79 individuals converted to dementia, whereas 119 remained with MCI status, see Table 3. In this dataset, atrophy/enlargement rate predictions were generated within a cross-validation framework. For MCI/dementia progression prediction, we selected results from the cross-validation run with the median PCC and used the predicted atrophy/enlargement rates from that run to predict MCI/dementia progression.

Second, we trained our models to predict atrophy/enlargement rates 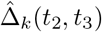 using the main ADNI dataset, and applied them to the AIBL cohort. The predicted rates in AIBL were then used to assess MCI/Dementia progression prediction. In the CN group, 13 individuals progressed from CN to MCI/dementia, while 45 remained stable. Due to the limited number of participants in the MCI group (*n* = 8) with available follow-up data extending five years beyond the current visit (i.e., the 18-month assessment), prediction of MCI-to-dementia progression was not conducted in the AIBL cohort. Lastly, we selected a subset of ADNI participants who were excluded from the main experiments due to missing 48-month data. However, since these individuals had both 0- and 24-month follow-up visits along with sufficient clinical follow-up information, they were eligible for inclusion in the progression prediction analysis. We refer to this subset as the ADNI-external test set. Within this subset, 25 individuals in the CN group progressed to MCI/dementia, while 56 remained stable, but the mean age of these two groups was highly different. In the MCI group, 46 individuals progressed to dementia, whereas 43 remained unchanged in their clinical status, see Table 3.

## 3 Results

### 3.1 Prediction of atrophy/enlargement: Cross-validation with ADNI

The validation results of all models are presented in Figure 2. Overall, our findings highlight that the longitudinal approach, particularly when incorporating additional risk factors, yielded the best performance for predicting annual atrophy/enlargement.

**Fig. 2.**
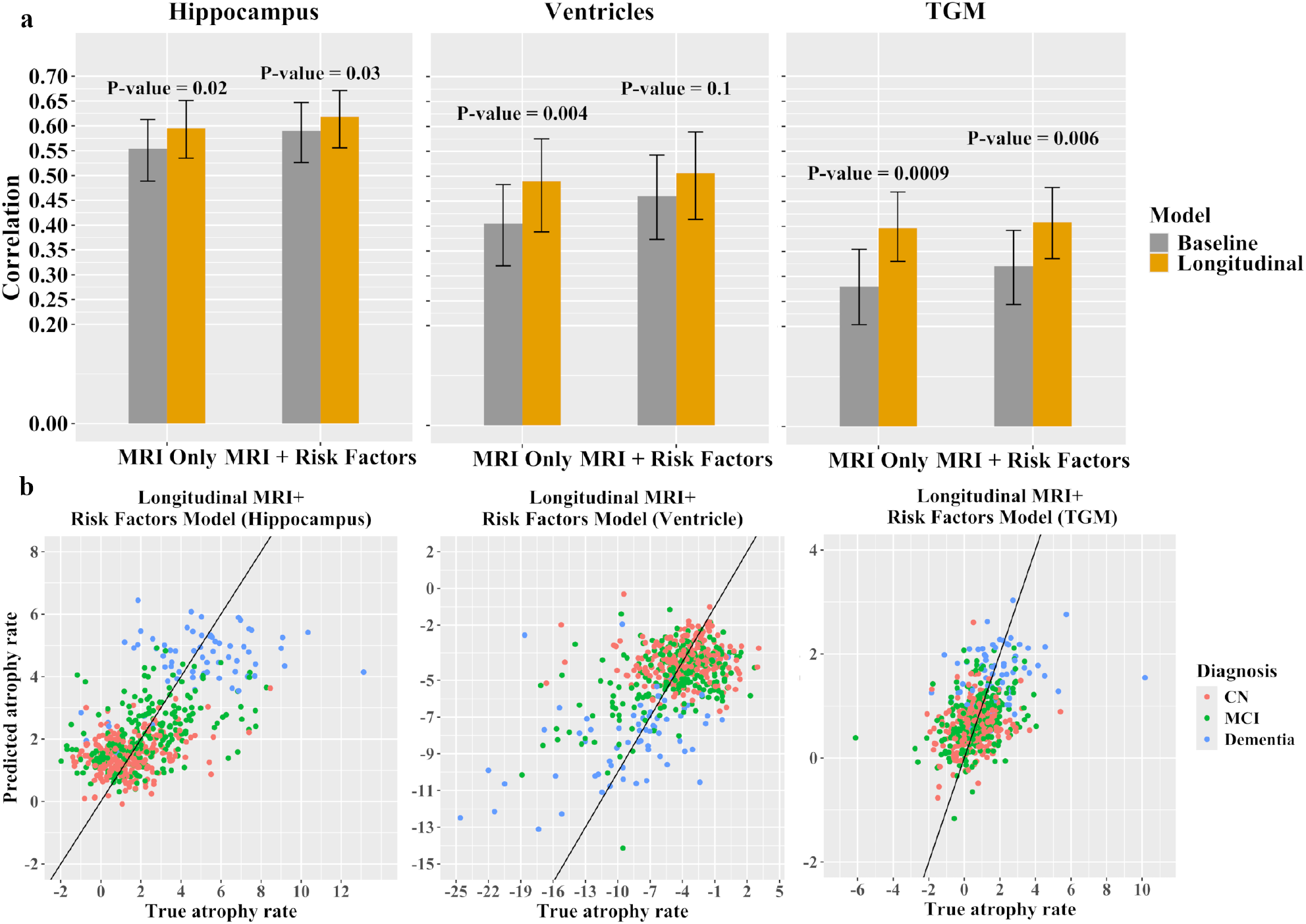
Prediction of annual percentage change of volumes of different brain regions using 10-fold cross-validation on the main ADNI dataset. **(a)** Bar plots showing the correlation of different models based on baseline and longitudinal approaches. **(b)** Scatter plot illustrating the prediction of atrophy rates for the longitudinal approach of MRI + Risk Factors model, derived from a single computation run with median performance.

#### Hippocampal atrophy

For the baseline approach utilizing only MRI data, the PCC for predicting the hippocampal atrophy rate was 0.55 (95% CI: 0.49, 0.61), with MAE of 1.36 (95% CI: 1.26, 1.46). Incorporating risk factors into the baseline MRI model, the PCC improved significantly to 0.59 (95% CI: 0.53, 0.65, p = 0.02), and the MAE decreased to 1.32 (95% CI: 1.22, 1.41). For the longitudinal approach using the MRI-only model, the PCC was 0.60 (95% CI: 0.54, 0.65), with an MAE of 1.31 (95% CI: 1.22, 1.41). When the longitudinal MRI model was combined with risk factors, the PCC significantly improved to 0.62 (95% CI: 0.56, 0.67, p = 0.04), with an estimated MAE of 1.29 (95% CI: 1.20, 1.38). In the MRI-only model, the PCC for the baseline approach significantly improved from 0.55 (95% CI: 0.49, 0.61) to 0.60 (95% CI: 0.54, 0.65) for the longitudinal approach (p = 0.02). Similarly, in the model incorporating both MRI and risk factors, the PCC increased from 0.59 (95% CI: 0.53, 0.65) in the baseline approach to 0.62 (95% CI: 0.56, 0.67) in the longitudinal approach, representing a significant improvement (p = 0.03).

#### Ventricle enlargement

For predicting ventricle enlargement rate using the baseline approach relying solely on MRI data, the PCC was 0.40 (95% CI: 0.32, 0.48), with an MAE of 2.67 (95% CI: 2.48, 2.88). Incorporating risk factors into the baseline MRI model improved the PCC significantly to 0.46 (95% CI: 0.37, 0.54, p = 0.004), while reducing the MAE to 2.57 (95% CI: 2.38, 2.79). For the longitudinal approach using the MRI-only model, PCC increased to 0.49 (95% CI: 0.39, 0.58), and the MAE decreased to 2.52 (95% CI: 2.33, 2.72). The best performance was achieved with the longitudinal approach combining the MRI model and additional risk factors, yielding the PCC 0.51 (95% CI: 0.41, 0.59) and the MAE of 2.48 (95% CI: 2.29, 2.69), however, this improvement was not significant (p = 0.4) over the longitudinal MRI-only model. In order to compare the models using only MRI information for ventricle enlargement rate, the PCC for the baseline approach was 0.40 (95% CI: 0.32, 0.48). For the longitudinal approach, the PCC increased to 0.49 (95% CI: 0.39, 0.58), representing a significant improvement (p = 0.004). For the model incorporating both MRI and risk factor information, there was no significant improvement, with the PCC increasing from 0.46 (95% CI: 0.37, 0.54) in the baseline approach to 0.51 (95% CI: 0.41, 0.59) in the longitudinal approach (p = 0.1).

#### Gray matter atrophy

To predict the TGM atrophy, based on baseline approach utilizing only MRI data, the PCC was 0.28 (95% CI: 0.20, 0.35), with a MAE of 0.90 (95% CI: 0.83, 0.97). Incorporating risk factors into the MRI model improved the PCC to 0.32 (95% CI: 0.24, 0.39) and slightly reduced the MAE to 0.89 (95% CI: 0.82, 0.95), the difference between the two models was not significant (p = 0.1). For the longitudinal approach using only the MRI model, the PCC increased further to 0.40 (95% CI: 0.33, 0.47), with the MAE of 0.86 (95% CI: 0.80, 0.93). The longitudinal MRI+Risk factors model achieved a PCC of 0.41 (95% CI: 0.34, 0.48) and an MAE of 0.86 (95% CI: 0.80, 0.93). The improvement over longitudinal MRI-only model was not significant (p = 0.6). In the MRI-only model, the PCC was 0.28 (95% CI: 0.20, 0.35) for the baseline approach and increased to 0.40 (95% CI: 0.33, 0.47) for the longitudinal approach, demonstrating a significant improvement (p= 0.0009). Furthermore, after incorporating risk factors alongside MRI information, we observed a significant improvement from 0.32 (95% CI: 0.24, 0.39) in the baseline approach to 0.41 (95% CI: 0.34, 0.48) in the longitudinal approach (p = 0.006).

#### Predictor importance

The most important predictors for atrophy/enlargement rates in the longitudinal MRI+risk factor model are displayed in Fig. 3. For predicting hippocampal atrophy, the five most important predictors were dementia diagnosis, right hippocampal volume, and changes in volumes of left middle temporal gyrus, right hippocampus and left cerebral white matter. Both present day volumes and changes in volumes of various brain structures contributed to models. However, only present day cortical thickness values (and not the rate of change in cortical thickness) were included as top predictors. From risk factors, only dementia diagnosis and the number of APOE4 alleles were included in the top predictors. Unsurprisingly, only the left or right (but not both) structure was included in the top predictors, with the exception of hippocampus volume. For predicting ventricle enlargement, the five most important predictors were changes in volumes of left lateral ventricle, left middle temporal gyrus, and left parahippocampal gyrus, dementia diagnosis, and the number of APOE4 alleles. The change rates of volumes of various structures appeared more often in the top predictors than the present day volumes of structures. Few cortical thickness measures were included as well as change rate of left rostral middle frontal cortex. Again, from risk factors, only dementia diagnosis and the number of APOE4 alleles were included in the top predictors. For TGM atrophy, the five most important predictors were dementia diagnosis, change rates of right parahippocampal gyrus volume, left transverse temporal cortical thickness, right cerebellum white matter volume, and left isthmus cortical thickness. Again, the change rates of volumes of various structures appeared more often in the top predictors than the present day volumes of structures. Both changes of and present day cortical thicknesses were included as top 20 predictors. The only risk factor in the top predictor list was the dementia diagnosis. For TGM, one of variables coding the MRI field strength entered to the model (strength3, indicating that all the measurements were done with 3.0T scanner).

**Fig. 3.**
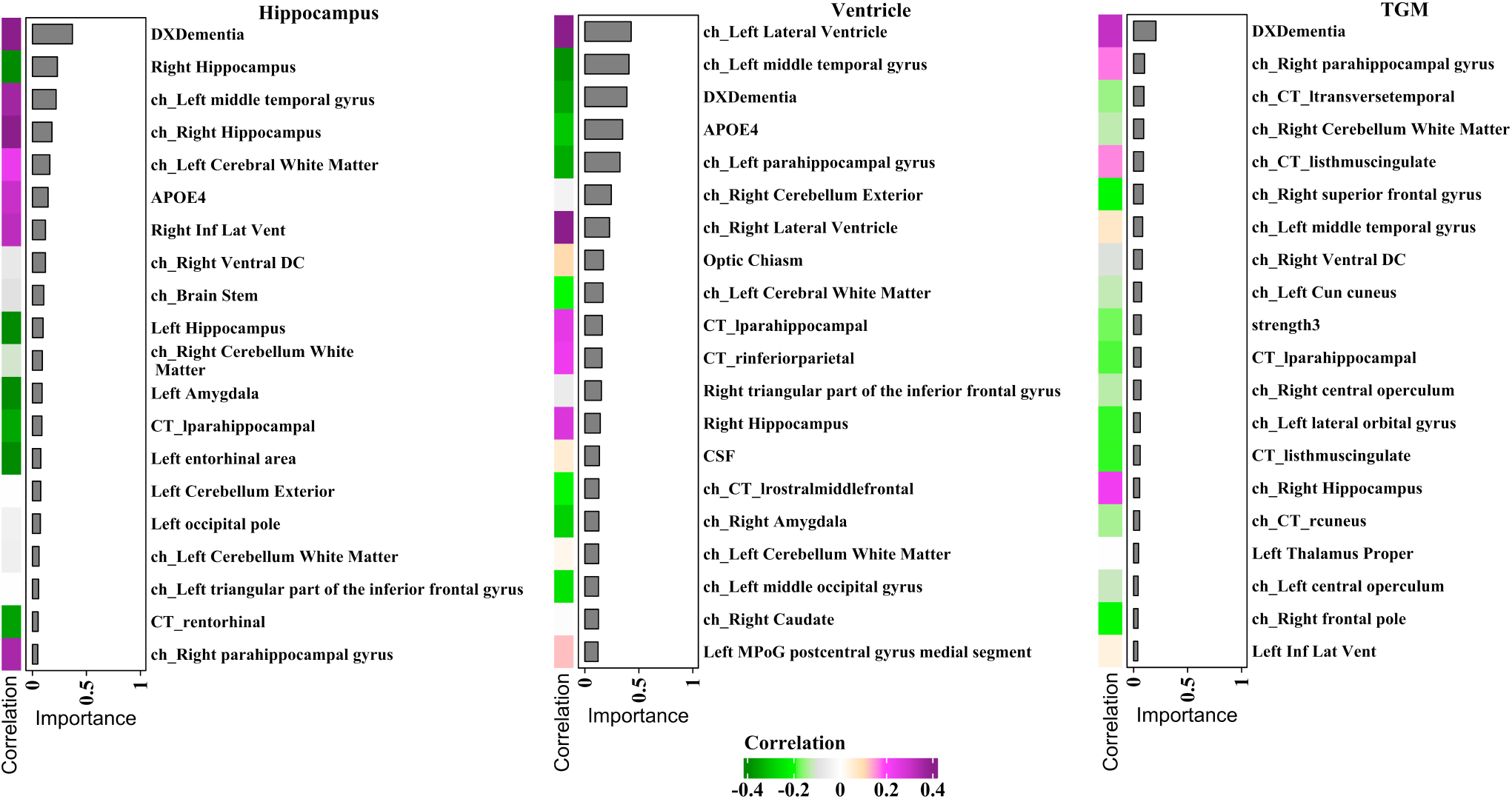
Importance of predictors across 10 runs of 10-fold CV for the longitudinal, MRI+Risk factors model. Each heatmap consists of a separate single-column heatmap showing the correlation scores between each variable and the target label. Additionally, a bar graph illustrates the importance of each predictor, calculated as the mean of absolute values of regression coefficients across the cross-validation folds. Note that the regression coefficients are computed for standardized models. For brain imaging predictors, “ch” indicates change and “CT” indicates cortical thickness. Predictor names without “CT” refer to volumes. “DX” refers to the diagnosis at 24 months, defined across three columns: “DX-CN”, “DX-Dementia”, and “DX-MCI”. Strength refers to variable coding of the field strengths of the scanners used to acquire the scans.

### 3.2 Prediction of atrophy/enlargement: External evaluation with AIBL

Figure 4 presents the external evaluation results for atrophy/enlargement prediction using the AIBL dataset. These results demonstrate that the longitudinal approach outperforms the baseline approach when tested on an external dataset.

**Fig. 4.**
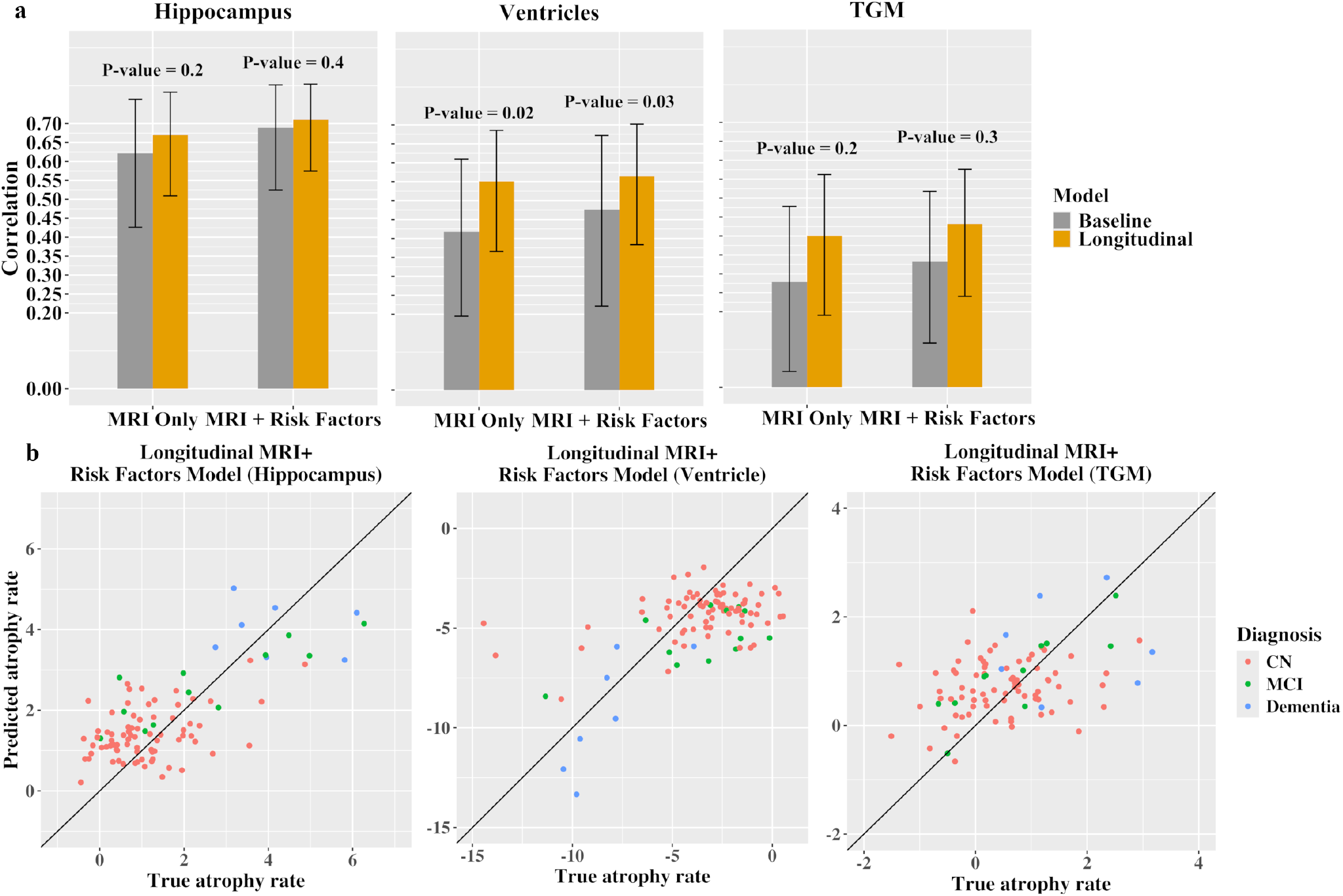
Prediction of annual percentage change of volumes in different brain regions on the AIBL (test) dataset. **(a)** Bar plots showing the correlation scores of different models based on baseline and longitudinal approaches. **(b)** Scatter plots illustrating the prediction of atrophy rates for the longitudinal approach of MRI + Risk Factors model, derived from a single computation run with median performance.

The external evaluation for hippocampal atrophy prediction using the baseline approach with the MRI only model resulted in a PCC of 0.62 (95% CI: 0.43, 0.76) and a MAE of 1.00 (95% CI: 0.87, 1.15). When applying the longitudinal approach and incorporating risk factors alongside MRI data, the PCC improved to 0.71 (95% CI: 0.57, 0.80), while the MAE decreased to 0.89 (95% CI: 0.78,1.01). The external evaluation for ventricle enlargement prediction using the baseline approach, which relied solely on MRI data, resulted in the PCC of 0.42 (95% CI: 0.19, 0.61) and the MAE of 2.40 (95% CI: 2.05, 2.79). In comparison, the longitudinal approach that combined MRI data with risk factor information achieved a higher PCC of 0.56 (95% CI: 0.38, 0.70) and a lower MAE of 2.14 (95% CI: 1.83, 2.46).

The external evaluation for TGM atrophy prediction using the baseline approach, which relied solely on MRI data, resulted in the PCC of 0.28 (95% CI: 0.04, 0.48) and the MAE of 0.82 (95% CI: 0.69,0.96). In comparison, the longitudinal approach that combined MRI data with risk factor information achieved a higher PCC of 0.43 (95% CI: 0.24, 0.58) and a slightly lower MAE of 0.78(95% CI: 0.66,0.90).

### 3.3 Prediction of MCI/dementia progression

The ROC curves assessing performance of the predicted atrophy/enlargement rates in clinical status progression prediction are presented in Figure 5 for ADNI main dataset and in Supplementary Figures 1 and 2 for AIBL and ADNI external datasets. All AUC statistics are collected in Table 4. Overall, the clinical status progression prediction results highlight that a) predicted atrophy yielded better than chance progression predictions and the predicted atrophy scores typically reached higher AUCs than the corresponding present day volume information.

**Table 4.**
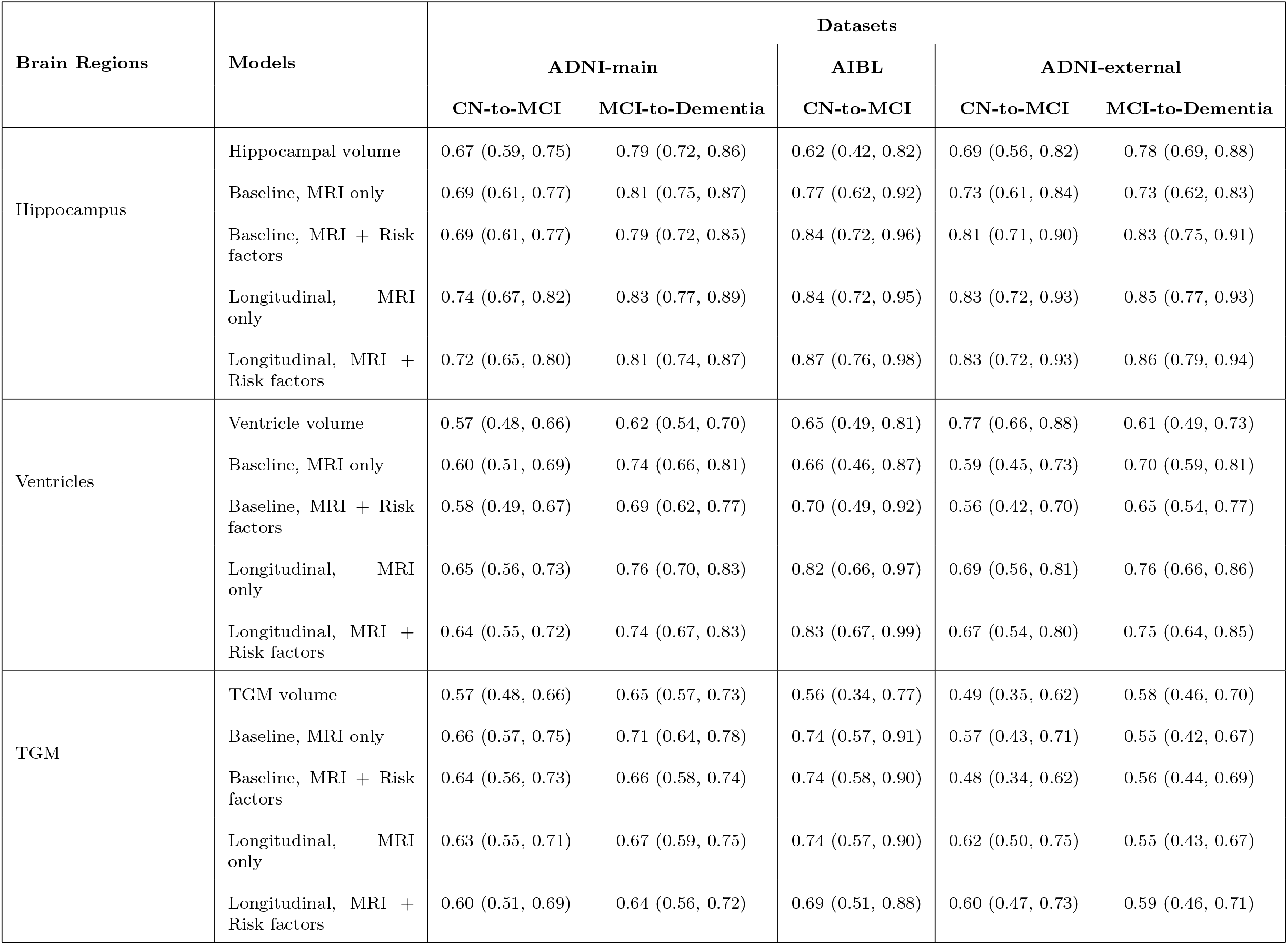
AUCs for different models for clinical status progression prediction across three databases. The values in parentheses refer to 95% confidence intervals of AUC.

**Fig. 5.**
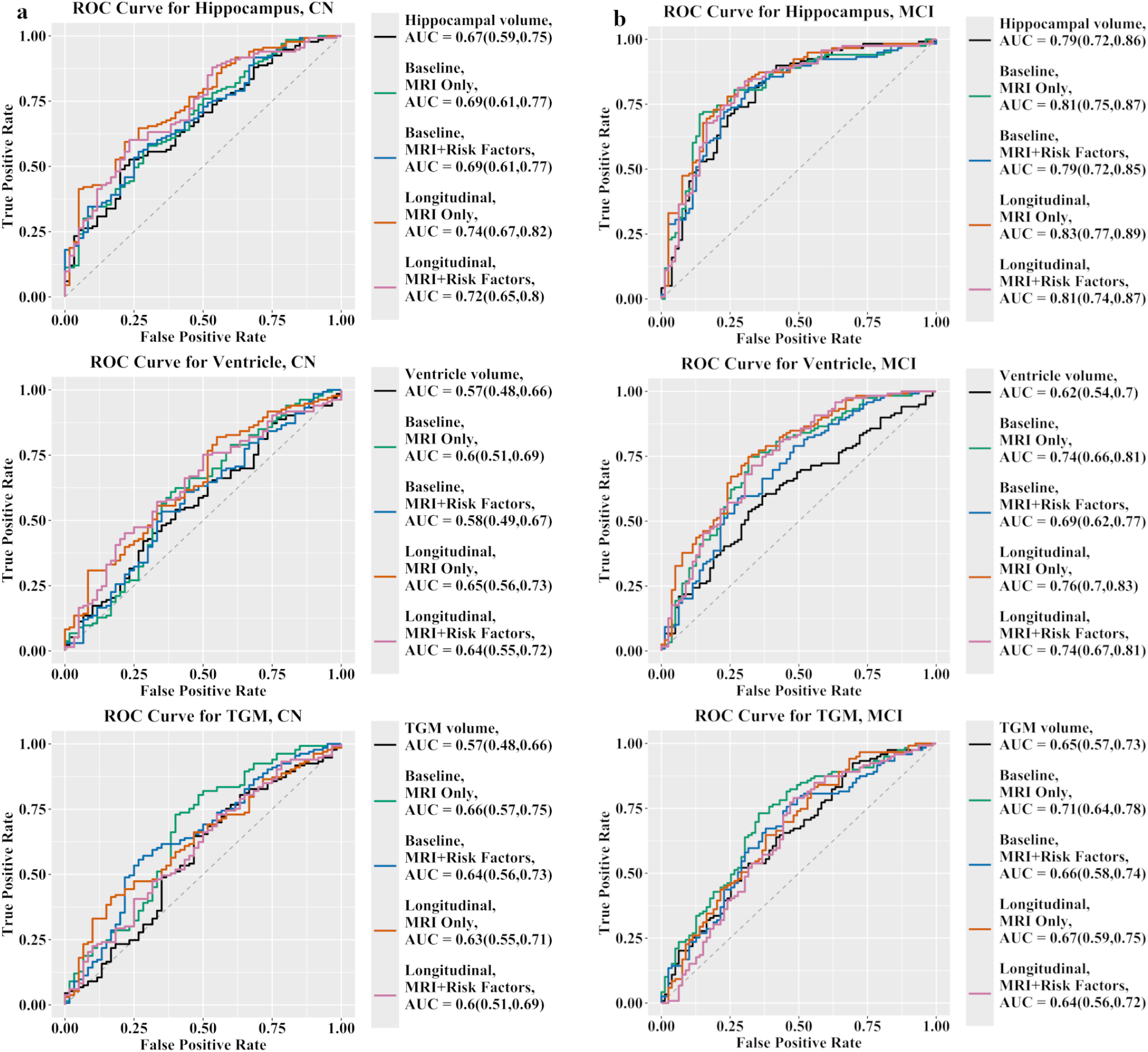
ROC curves of clinical status progression prediction in **a** CN and **b** MCI groups with different models and brain regions using ADNI-Main dataset.

#### ADNI-Main Cohort

For the ADNI-Main cohort, using present day hippocampus volume achieved an AUC of 0.67 (95% CI: 0.59, 0.75) for the CN-to-MCI progression prediction. The predicted hippocampal atrophy (with longitudinal MRI + risk factors model) resulted in the improved progression prediction performance, yielding an AUC of 0.72 (95% CI: 0.65, 0.80). However, this improvement was not statistically significant (p = 0.10, DeLong test [50]). Similarly, the present day ventricle volume produced an AUC of 0.57 for CN-to-MCI progression prediction (95% CI:0.48, 0.66), which increased to 0.64 (95% CI:0.55,0.72) with the predicted ventricle enlargement (longitudinal MRI + risk factors model). This improvement was not statistically significant (p = 0.30, DeLong test). The present day TGM volume predicted CN-to-MCI progression with an AUC of 0.57 (95% CI: 0.48, 0.66), which increased to 0.60 (95% CI: 0.51, 0.69) with the predicted TGM atrophy (longitudinal MRI +risk factors). This increment was not statistically significant (p = 0.60).

Within MCI group, present day hippocampus volume predicted progression to dementia with an AUC of 0.79 (95% CI: 0.72, 0.86). Predicted atrophy (longitudinal MRI + risk factors model) did not significantly improve the performance (p = 0.50), yielding an AUC of 0.81 (95% CI: 0.74, 0.87). For ventricle volume, the AUC was 0.62 (95% CI: 0.54, 0.70), which significantly improved to 0.74 (95% CI: 0.67, 0.81) when predicting progression to dementia using predicted atrophy (longitudinal MRI + risk factors model, p = 0.04). For the TGM volume, the AUC for the TGM volume was 0.65 (95% CI: 0.57, 0.73) and predicted atrophy for longitudinal MRI + risk factors model yielded a similar AUC of 0.64 (95% CI:0.56, 0.72) (p=0.9).

The comparison between different atrophy prediction models (baseline vs. longitudinal, MRI only vs. MRI + risk factors) did not yield any clearly best model, but all the models performed similarly in the progression prediction.

#### ADNI-External Cohort

For the ADNI-external cohort among the CN group, present day hippocampus volume achieved an AUC of 0.69 (95% CI: 0.56, 0.82) for the CN-to-MCI progression prediction. The predicted hippocampal atrophy (with longitudinal MRI + risk factors model) resulted in the increased progression prediction performance, yielding an AUC of 0.83 (95% CI: 0.72,0.93). This improvement was statistically significant (p = 0.04, DeLong test). Similarly, the present day ventricle volume produced an AUC of 0.77 for CN-to-MCI progression prediction (95% CI:0.66, 0.88), which reduced to 0.67 (95% CI:0.54,0.80) with the predicted ventricle enlargement (longitudinal MRI + risk factors model). This reduction was not statistically significant (p = 0.2, DeLong test) and we hypothesize that the high AUC for ventricle volume based prediction was driven by a large age difference between the stable and progressive groups (see Table 3). The present day TGM volume predicted CN-to-MCI progression with an AUC of 0.49 (95% CI: 0.35, 0.62), which increased to 0.60 (95% CI: 0.47, 0.73) with the predicted TGM atrophy (longitudinal MRI +risk factors). This increment was not statistically significant (p = 0.2).

For the MCI group in the ADNI-external, the model using hippocampal volume alone achieved an AUC of 0.78 (95% CI: 0.69, 0.88). With the predicted hippocampal atrophy (longitudinal MRI + risk factors model) the progression prediction performance improved significantly, yielding an AUC of 0.86 (95% CI: 0.79, 0.94) (p=0.03). The ventricle volume model achieved an AUC of 0.61 (95% CI: 0.49,0.73), which increased to 0.75 (95% CI: 0.64,0.85) with the predicted ventricle enlargement (longitudinal MRI + risk factors mode), but the increase was not significant (p = 0.1). The model using TGM volume resulted in an AUC of 0.58 (95% CI: 0.46,0.70), while the predicted TGM atrophy (longitudinal MRI + risk factors model) resulted to a similar AUC to 0.59 (95% CI: 0.46, 0.71) (p = 0.9).

Notably, the AUCs for clinical status progression prediction were consistently higher for atrophy scores predicted based on longitudinal model than for the atrophy scores based on baseline model. Addition of risk factors to the atrophy score prediction did not have much influence on the progression prediction, with the exception of the hippocampus and the baseline model, where atrophy scores predicted based on MRI + risk factors model provided better progression predictions than those based on MRI only model.

#### AIBL Cohort

Using hippocampus volume, we could predict the progression from CN to MCI with an AUC of 0.62 (95% CI: 0.42, 0.82). Using predicted atrophy scores significantly improved the prediction performance, resulting in an AUC of 0.87 (95% CI: 0.76, 0.98) (p=0.02), highlighting the enhanced predictive value of this approach. The ventricle volume achieved an AUC of 0.65 (95% CI: 0.49, 0.81) while the AUC with predicted ventricle enlargement improved to 0.83 (95% CI: 0.67, 0.99), however, the improvement was not significant (p = 0.1). Using TGM volume, the AUC was 0.56 (95% CI: 0.34, 0.77) while the AUC with predicted TGM was increased to 0.69 (95% CI: 0.51, 0.88) (p = 0.3).

Here, the longitudinal MRI + risk factors model was clearly superior to the baseline MRI only model for progression prediction using predicted hippocampal atrophy and ventricle enlargement. With TGM, all the four models performed similarly in terms of progression prediction.

## 4 Discussion

Hippocampal atrophy, ventricle enlargement, and whole-brain atrophy are three key MRI markers used to predict AD/dementia [16, 51, 52]. This study aimed to predict the annual change in the hippocampus, ventricles, and total gray matter volumes (used as a proxy for whole-brain atrophy), in individuals with varying levels of cognitive impairment, ranging from healthy to dementia. To achieve this, we developed an ML model based on ENLR. We explored two approaches: (1) a baseline method relying on single-time-point data and (2) a longitudinal method incorporating information from two years prior to baseline. Both approaches were tested using models that utilized only MRI data and models that combined MRI data with additional risk factors, including age, sex, and baseline diagnosis. Our analysis also explored how incorporating longitudinal data affected the performance of these models.

The predictions for all three MRI markers were clearly better than the baseline prediction of no chance that would have the PCC of zero. Based on the PCC values, the hippocampal atrophy could be best predicted, followed by the ventricle enlargement, while the TGM atrophy was the most challenging to predict. Indeed, with the external test data (AIBL) hippocampal atrophy prediction had PCC of 0.71 while TGM atrophy prediction only reached PCC of 0.43.

Our findings showed that incorporating additional information into the baseline MRI model significantly enhanced its ability to predict hippocampal atrophy and ventricular enlargement. In our previous works, we have demonstrated the advantages of combining multiple data modalities—such as MRI and cognitive assessments — for prediction tasks relevant to dementia management[27, 36, 53]. Also, other studies have emphasized the importance of integrating diverse data sources for enhanced predictive accuracy[25, 54, 55]. Our current results align with these findings, but with a notable difference in the methodology. Instead of using baseline cognitive assessments as predictors, we utilized baseline diagnoses as predictors. Baseline diagnosis serves as a summary of cognitive assessments, encapsulating an individual’s cognitive status. Using a single summary variable reduced data dimensionality and redundancy, as many cognitive measures convey overlapping information.

Incorporating data from two years prior to the current visit into a longitudinal model consistently enhanced predictive performance even if the performance improvement was in relatively modest especially in hippocampal atrophy prediction. With ADNI, the improvement was statistically significant when comparing baseline versus longitudinal using MRI-only models across all cases, including hippocampal atrophy, TGM atrophy, and ventricular enlargement. For MRI + risk factor models, adding historical data also improved performance, but the improvement was smaller compared to the MRI-only models. The improvement remained statistically significant for hippocampal and TGM atrophy but not for ventricular enlargement. Overall, the best predictions were achieved using the longitudinal MRI + risk factor model across all three brain regions. External validation using the AIBL dataset confirmed these findings, demonstrating that longitudinal models consistently provided more accurate predictions for all three types of annual change. Our findings extend those of earlier studies that have shown the importance of considering longitudinal data in prediction tasks based on brain imaging data [56, 57]. We speculate that the greater improvement observed in the MRI-only model compared to the MRI + risk factor model may be due to the absence of cognitive status information in MRI-only models. Adding historical data to baseline MRI data provides additional insights into individual structural changes over time, as the differences between past and baseline MRI scans vary across diagnostic groups (CN, MCI, AD). This temporal information helps the model differentiate individuals more effectively. In contrast, the MRI + risk factor model already includes diagnostic group information, which partially accounts for cognitive status, thereby reducing the relative contribution of historical data.

In longitudinal models, we investigated the importance of various predictors, including risk factors, MRI features and their annualized percentage changes between baseline and two years prior. A closer examination of the coefficient values in the longitudinal model revealed that baseline diagnosis is one of the most important features for predicting the annual percentage change in all three studied markers. Among the top selected features, APOE4 emerged as a significant predictor for hippocampal atrophy and ventricular enlargement, but not for TGM atrophy. Interestingly, many longitudinal MRI features were identified as important within the top selected features, emphasizing the value of incorporating longitudinal information into the models. Another notable observation is that, for predicting the annual percentage change in each biomarker, multiple brain regions contribute to the prediction task, rather than the respective region alone holding the highest importance. Among the top selected MRI features for predicting hippocampal atrophy and ventricular enlargement were the hippocampus, middle temporal gyrus, inferior lateral ventricles, and amygdala, all of which are highly relevant in AD progression [51]. However, the top selected features for predicting TGM atrophy differed slightly from those for hippocampal and ventricular predictions and were not as strongly associated with AD. These results align with our expectations, as the hippocampus and ventricles are the primary brain regions affected by AD, even in the early stages of disease. In contrast, TGM represents the whole brain and exhibits a weaker association with AD compared to the hippocampus and ventricles. Overall, our findings indicate that multiple brain regions, along with their longitudinal information and associated risk factors, play a crucial role in predicting the annual percentage change in all these biomarkers.

The MRI scans in the ADNI dataset were drawn from different ADNI phases, and MRI data was acquired using different field strengths. This variability posed a challenge [58, 59], particularly as our analysis considered multiple time points for each participant, two time points (current day and future) for baseline models, and three time points (past, current day, and future) for longitudinal models. For many participants, the field strength was not consistent across all time points. To address this issue, we introduced a new categorical variable representing the field strength at each time point and included it as a predictor in our models (see [35]). By doing so, we accounted for the variability in field strength and ensured it was considered in the analysis, reducing its potential impact on the model’s performance. To streamline our analysis, we opted for a cross-sectional pipeline for MRI preprocessing using CAT12, prioritizing its simplicity and efficiency in reducing processing time. Our preliminary analyses revealed no significant performance differences between the cross-sectional and longitudinal preprocessing approaches for atrophy prediction in CAT12. Consequently, the cross-sectional pipeline was selected as a practical and effective choice for our study and the finding that longitudinal predictors outperformed cross-sectional ones supports this choice. However, the optimal approach for addressing the construction of the longitudinal predictors should be addressed systematically in future studies.

Although hippocampal and TGM atrophy, as well as ventricular enlargement, are critical factors in dementia progression, predicting volume changes in these regions has not been extensively studied. In particular, limited research has evaluated all three regions within the same dataset and pipeline to assess their roles in dementia progression. While various studies have investigated the associations of these biomarkers with progression to dementia [52, 60, 61], the prediction of changes over time and their relationship with progression to MCI or dementia remains underexplored. The prediction of total ventricular volume has been previously examined in the TADPOLE challenge [10], where the tasks were predicting clinical diagnosis, ADAS-Cog13 scores, and total ventricular volume. Another study by Liedes et al. [25] focused on predicting the annualized rate of hippocampal atrophy using multivariate ML algorithms on the ADNI dataset. Our study shares similarities with Liedes et al [25] in predicting annualized hippocampal atrophy but differs in three key ways: (1) we analyzed all three regions—hippocampus, TGM, and ventricles—together within the same dataset and workflow, rather than focusing solely on hippocampal atrophy; (2) we developed a longitudinal model that incorporates data from prior time points, enabling us to investigate the impact of longitudinal predictors on the performance of our predictive models; and (3) we evaluated the ability of our models to predict progression to MCI or dementia in both healthy and MCI groups.

We assessed the effectiveness of our predictive models in estimating the annual percentage changes in hippocampal, TGM, and ventricular volumes to predict dementia progression in both healthy and MCI groups. Results from the main ADNI dataset showed that predicted hippocampal atrophy and ventricular enlargement outperformed actual biomarker volumes in predicting progression to MCI or dementia in both CN and MCI groups. The most notable improvements came from predicted hippocampal atrophy, particularly using longitudinal approaches. For CN individuals, the predictive accuracy for progression to MCI/dementia improved from an AUC of 0.67 (using actual hippocampal volume) to 0.73 (using predicted atrophy with the longitudinal method). Similarly, in MCI individuals, the AUC for predicting progression to dementia increased from 0.79 to 0.83. However, no such improvement was observed for TGM atrophy, as predicted TGM atrophy did not provide better predictions compared to actual TGM volume and AUCs remained mostly below 0.70. Results from the external validation datasets, (AIBL and ADNI-external) showed similar trends. While our models were not specifically designed to predict progression to MCI or dementia, the predicted annual percentage changes—especially for hippocampal atrophy—performed well and the performance was comparable to previous MRI-based approaches for predicting dementia progression in CN individuals [62, 63]. This analysis demonstrates that predicted future atrophy scores can function as single valued and easily interpretable indexes of potential future cognitive decline. Also, they might be applicable when selecting/stratifying population for clinical trials, particularly when combined with disease-specific biomarkers.

In summary, our study highlights the importance of incorporating longitudinal data to improve predictions of annual changes in key biomarkers linked to dementia. By integrating MRI data and risk factors into machine learning models, we significantly enhanced predictive accuracy, particularly for hippocampal atrophy and ventricular enlargement. The inclusion of longitudinal data allowed us to capture individual structural changes over time, improving the model’s ability to differentiate between diagnostic groups. These findings emphasize the advantages of combining diverse data sources and tracks brain changes over time to better predict dementia progression. Future research should further explore the integration of multiple data modalities and assess the long-term impact of longitudinal data to refine predictive models for dementia.

## Supporting information

Supplementary.pdf (Corrected Results)

## Data Availability

All data produced are available online at
http://adni.loni.usc.edu
https://aibl.org.au

## Availability of data and materials

Data used in the preparation of this article were obtained from the Alzheimer’s Disease Neuroimaging Initiative (ADNI) database (https://adni.loni.usc.edu/) and and the Australian Imaging Biomarkers and Lifestyle flagship study of ageing (AIBL) (https://aibl.csiro.au/adni/index.html). Details about data access are detailed there. The participant’s RIDs are provided in Supplementary Materials. The codes to reproduce the work are available at https://github.com/MaryamHadji/Future-prediction. CAT12 toolbox is available at https://neuro-jena.github.io/cat.

## Competing interests

The authors have no actual or potential conflicts of interest.

## Funding

This research has been supported by The Academy of Finland, grant 351849 under the frame of ERA PerMed (“Pattern-Cog”), grants 346934 (PRIMAL), and 358944 (Flagship of Advanced Mathematics for Sensing Imaging and Modeling) from the Research Council of Finland.

## Author Contributions

M.H: Methodology, formal analysis, investigation, software, visualization, writing—original draft, and writing—review and editing; E.M: Methodology, formal analysis, software, writing—original draft, and writing—review and editing; J.T: Conceptualization, methodology, resources, funding acquisition, writing—original draft, and writing—review and editing.

## Acknowledgments

Data used in the preparation of this article were obtained from the Alzheimer’s Disease Neuroimaging Initiative (ADNI) database (www.adni.loni.usc.edu) and the Australian Imaging Biomarkers and Lifestyle flagship study of ageing (AIBL) funded by the Commonwealth Scientific and Industrial Research Organisation (CSIRO), which was made available at the ADNI database. The investigators within ADNI and AIBL contributed to the design and implementation of ADNI/AIBL and/or provided data but did not participate in analysis or writing of this report. A complete listing of ADNI investigators can be found at http://adni.loni.usc.edu/wp-content/uploads/how_to_apply/ADNI_Acknowledgement_List.pdf. AIBL researchers are listed at www.aibl.csiro.au.

Data collection and sharing for this project was funded by the Alzheimer’s Disease Neuroimaging Initiative (ADNI) (National Institutes of Health Grant U01 AG024904) and DOD ADNI (Department of Defense award number W81XWH-12-2-0012). ADNI is funded by the National Institute on Aging, the National Institute of Biomedical Imaging and Bioengineering, and through generous contributions from the following: AbbVie, Alzheimer’s Association; Alzheimer’s Drug Discovery Foundation; Araclon Biotech; BioClinica, Inc.; Biogen; Bristol-Myers Squibb Company; CereSpir, Inc.; Cogstate; Eisai Inc.; Elan Pharmaceuticals, Inc.; Eli Lilly and Company; EuroImmun; F. Hoffmann-La Roche Ltd and its affiliated company Genentech, Inc.; Fujirebio; GE Healthcare; IXICO Ltd.; Janssen Alzheimer Immunotherapy Research & Development, LLC.; Johnson & Johnson Pharmaceutical Research & Development LLC.; Lumosity; Lundbeck; Merck & Co., Inc.; Meso Scale Diagnostics, LLC.; NeuroRx Research; Neurotrack Technologies; Novartis Pharmaceuticals Corporation; Pfizer Inc.; Piramal Imaging; Servier; Takeda Pharmaceutical Company; and Transition Therapeutics. The Canadian Institutes of Health Research is providing funds to support ADNI clinical sites in Canada. Private sector contributions are facilitated by the Foundation for the National Institutes of Health (www.fnih.org). The grantee organization is the Northern California Institute for Research and Education, and the study is coordinated by the Alzheimer’s Therapeutic Research Institute at the University of Southern California. ADNI data are disseminated by the Laboratory for Neuro Imaging at the University of Southern California.

During the preparation of this work, the authors used GPT-4 from OpenAI and Microsoft Copilot to improve language and readability. After using this tool/service, the authors reviewed and edited the content as needed. The authors take full responsibility for the content of the publication.

The computational analyses were partly performed on servers provided by the UEF Bioinformatics Center and Biocenter Kuopio, Biocenter Finland, University of Eastern Finland, Finland.

## Footnotes

1 The atlas is derived from the maximum probability tissue labels derived from the “MICCAI 2012 Grand Challenge and Workshop on Multi-Atlas Labeling” http://www.neuromorphometrics.com/2012 MICCAI Challenge Data.htm with the MRIs from OASIS project and the labeled data provided by Neuromorphometrics, Inc. (Neuromorphometrics.com) under academic subscription.

## Notes

### Competing Interest Statement

The authors have declared no competing interest.

### Summary of Updates

This version of the manuscript has been revised to incorporate several corrections and updates. Specifically, we made minor adjustments in the Results section to ensure consistency with the data analysis. The numerical values and descriptions have been checked for accuracy and clarity. In addition, the supplementary material has been updated to include the corrected site-specific results and corresponding tables. Minor editorial edits were also made throughout the text to improve readability and alignment with the revised findings.

