## Supplementary.pdf (Corrected Results) for "Predicting Future Brain Atrophy Based on Longitudinal MRI"

September 30, 2025

### Supplementary Tables and Figures

| **Brain region** | **Without harmonization** | | **Neuro Combat** | |
|---|---|---|---|---|
| | **Pearson, 95% CI** | **MAE, 95% CI** | **Pearson, 95% CI** | **MAE, 95% CI** |
| Hippocampus | 0.55 (0.49, 0.61) | 1.36 (1.25, 1.46) | 0.46 (0.40, 0.53) | 1.56 (1.45, 1.68) |
| Ventricles | 0.40 (0.32, 0.48) | 2.67 (2.48, 2.90) | 0.37 (0.29, 0.45) | 3.04 (2.80, 3.30) |
| TGM | 0.28 (0.20, 0.35) | 0.90 (0.83, 0.97) | 0.17 (0.10, 0.25) | 0.97 (0.90, 1.05) |

Table S1: Model performance with and without NeuroCombat harmonization of 1.5 T and 3.0 T MRI using the ADNI-Main dataset. The harmonization was performed on the measure-level (i.e., CAT12-derived regional measures were harmonized). The experimental settings were the same as reported in the main article. We compared only the results with the baseline MRI-only model. The performance was equal to or slightly worse after harmonization, which could be due to the fact that NeuroCombat is not designed for longitudinal settings such as in here.

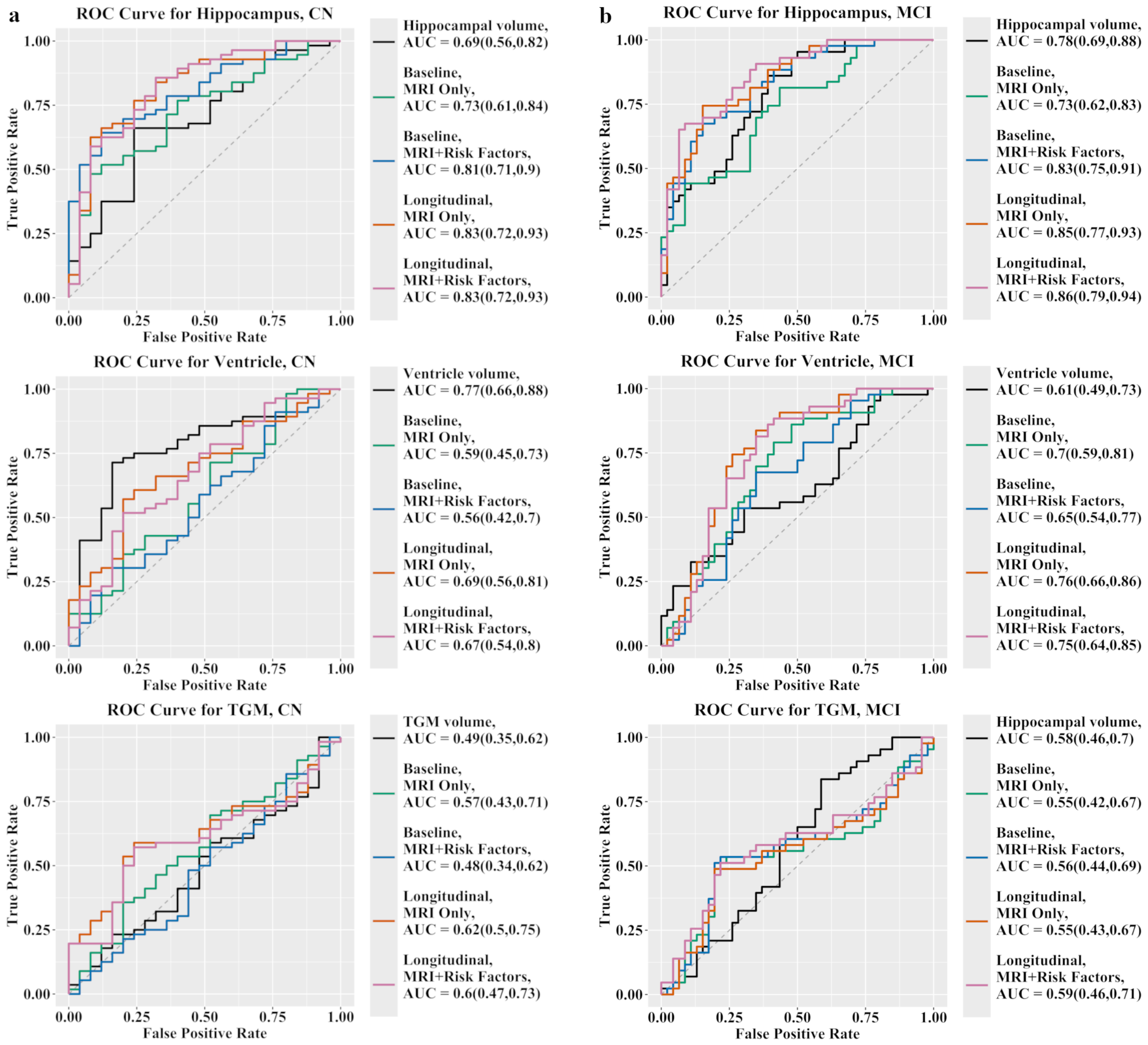


Figure S1: ROC curves of clinical status progression prediction in **a** CN and **b** MCI groups with different models and brain regions using ADNI-External dataset. The atrophy predictions were trained using ADNI-Main dataset.

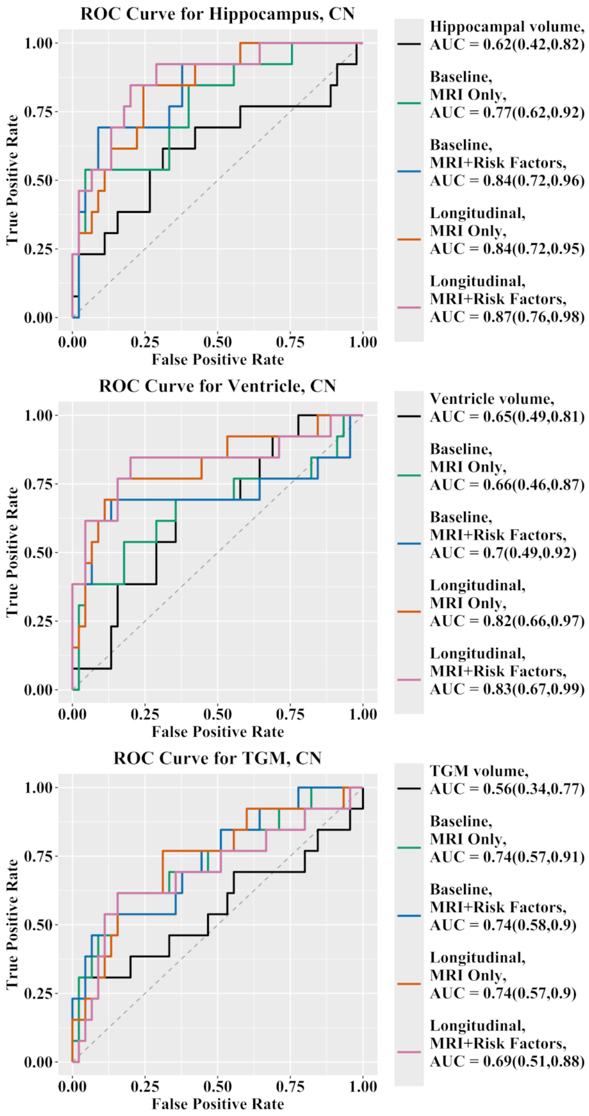


Figure S2: ROC curves of clinical status progression prediction in CN and MCI groups with different models and brain regions using AIBL dataset. The atrophy predictions were trained using ADNI-Main dataset.
